# Evaluating the Effectiveness of a Population-Level Health Intervention to Increment HCV Treatment Coverage in Tuscany Region, Italy: An Interrupted Time Series Analysis

**DOI:** 10.1101/2024.06.25.24309463

**Authors:** Chiara Seghieri, Luca Ceccarelli, Costanza Tortù, Lara Tavoschi

## Abstract

Worldwide, an estimated 71.1 million people are chronically infected with the Hepatitis C virus (HCV). The advent of direct-acting antivirals (DAAs) has made possible the definition of elimination targets by 2030. This study aimed to evaluate the effectiveness of a population-level health intervention to expand access to HCV treatment in the Tuscany Region, Italy.

We used individual-level administrative data from the Tuscany region, collected between January 2015 and December 2022. Data include monthly observations on i) the number of serological tests to detect HCV, ii) the number of PCR tests to detect HCV and, iii) the number of prescriptions of direct-acting antivirals against HCV.

We implemented an Interrupted Time Series (ITS) model, where the primary outcome was the number of monthly prescriptions of direct-acting antivirals, while the number of tests to detect HCV were included as control variables. The analysis was implemented i) in the general population, ii) in specific sub-population groups.

Results show that the health intervention promoted by the Tuscany Regional Health Authority was highly effective in increasing DAAs treatment coverage in the general population, while no significant effects were observed among sub-population groups.

Findings of this study provide evidence to support policies at national and subnational levels to booster HCV screening and simplify access to DAA prescriptions.

## INTRODUCTION

Acute and chronic hepatitis can be brought on by the hepatitis C virus (HCV), which increases the risk of cirrhosis, liver cancer, and death^1–2^. An estimated 3.9 million people in the European Union/Economic European Area (EU/EEA) are among the 57.8 million people with HCV^3^ who are chronically infected worldwide. Due to the fact that chronic HCV infection is frequently asymptomatic, a significant number of infected individuals go undiagnosed^4^. The prevalence of HCV is unequally distributed throughout the population, with people living in prison (PLP), people who use drugs (PWUD), people living withHIV-positive (PLHIV), or people with certain chronic illnesses (e.g. patients undergoing hemodialysis, diabetics, or those who received blood or other products of human origin) being disproportionately affected^5^. At present, no studies have evaluated the effectiveness of population-level health policies implemented to increment HCV test and treatment coverages in Italy.

According to the available evidence^6–11^, an estimated 398,610 people in Italy (1.7% of the population) have active HCV infection, with central regions having the highest prevalence (0.88%), followed by southern and island regions (0.72% and 0.67%), and northern regions (0.54%)^12–13^. Existing surveillance for viral hepatitis in Italy only monitors acute cases^8^, thus limiting the system’s ability to assess the burden of both acute asymptomatic and chronic illness. Large clinical cohort studies were established to foster research initiatives, primarily on clinical grounds^12^. Epidemiological research using administrative health information and data linkage approaches have been rarely used to overcome the lack of data. A data-linkage study was carried out in Tuscany, a region with more than 3 million inhabitants, to estimate the prevalence of people with chronic HCV infection and in need of treatment by the end of 2015^14^. This and other seroprevalence studies conducted in Italy^15^, concur to an estimated prevalence of 1%.

The HCV elimination targets by 2030, including a decrease in the percentage of undiagnosed cases and an increase in treatment coverage, were made achievable by the development of direct-acting antivirals (DAA). The World Health Organization (WHO) defined numerous targets along the continuum of care for HCV to help reach this goal, including the diagnosis and treatment of 90% of persons with chronic HCV infection by 2030^16^. The European Office of the WHO has developed a Regional Action Plan that prioritizes efforts to combat disease and adapt global targets to the regional epidemiological context^17^. The Italian National Plan for the Prevention of Viral Hepatitis (PNEV), approved by the Italian Ministry of Health in 2015^18^, included the global and regional objectives for HCV eradication.

Since 2014, patients with severe diseases in Italy have been entitled to receive free therapy at the point of care upon a specialist’s prescription. Through a monitoring registry for DAAs, the Italian Medicines Agency (AIFA) keeps track of new patients who start receiving medication (https://www.aifa.gov.it/aggiornamento-epatite-c). With the 2017 AIFA resolution, expanded access to DAA has been achieved in Italy^19^ and a national registry of patients initiating treatment was established and updated regularly. At regional level, in Tuscany a triennial regional action plan was launched in 2018 with the aim of raising the treatment coverage and contributing to the elimination of HCV in the region^20^. The plan foresaw more than 6,000 HCV chronically infected individuals to be treated every year up to 2020. Yet, identification of infected individuals and linkage to care posed a challenge to the achievement of elimination targets. A national one-time screening program^23^ was launched at the end of 2021 with a focus on PLP, PWUD, and the birth cohorts from 1969 to 1989, approximately corresponding to the people aged between 33 and 53.

Given these premises, the goal of this study is twofold: a) to evaluate the effectiveness of the population-level health interventions implemented since 2018 in Tuscany to expand access to HCV treatment b) to evaluate the impact of COVID-19 pandemic on the regional action plan. Interrupted time series analysis was applied on the monthly time series health administrative data to evaluate the effect of the regional plan. Study results provide valuable evidence for decision-makers including recommendations on further interventions that may be needed.

## MATERIAL AND METHODS

### Tuscany Regional Healthcare System

The National Health Service (NHS) is the national healthcare system in Italy. There are three tiers to the system organization: local, regional, and national. The national level is in charge of defining the overarching goals and core values of the NHS. The two Autonomous Provinces (R&AP) and the 19 regions are in charge of planning and executing the provision of healthcare. Local health authorities (LHAs) in each region are responsible for providing primary care, public health services, community health services, and specialized treatment either directly or through public hospitals or private providers with accreditation. Each LHA’s geographic area is further subdivided into Health Care Districts (HCD), which are in charge of offering primary care and public health services^.24^.

The Italian health care system provides free care to every citizen who has a specific code for exemption from healthcare costs (exemption code). Exemption codes may be given to individuals after the diagnosis of a specific disease (e.g. HCV) or because of a particular socio-medical condition requiring tailored care and treatments (e.g. problematic drug use, imprisonment).

Tuscany is a region of central Italy that counts 3,673,347 inhabitants with three LHA (North-West, Center, South-East) and four university hospitals, to which 12 territories (provinces) refer. In Tuscany, about 90% of healthcare services are delivered by entities that are either public providers or private accredited providers. In the region there are 14 prisons, 40 harm reduction services (SerD) and 16 recognized non-governmental organizations (NGOs) that provide services to PWUD. The LHA (Ausl), 3 in Tuscany region, are territorial branches of the regional health service and guarantee the homogeneity of assistance in the different areas of the region. Divided into district-areas, they provide for the management and planning of the activities defined in the uniform and essential levels of care, including socio-medical services with high health integration, health services of social relevance and social assistance activities delegated to local authorities.

### Sources of data

We used individual-level administrative data from the Tuscany region, collected between January 2015 and December 2022. The system can track each healthcare service, identified in the system by a unique code, provided to each individual, identified in the system by a unique personal identifier (alphanumeric code). However, outcomes of the services provided (e.g., exams’ results) were not available in the system in line with current national and regional privacy regulations.

All data recorded in the Tuscan healthcare administrative databases are pseudo-anonymized at the Regional Health Information System Office whereby each individual is assigned a unique identifier. The study was carried out in compliance and accordance with the General Data Protection Regulation (2016/679) and the Italian Legislative Decree No. 196/2003 (“Personal Data Protection Code ”) within the project OPT-HepaC - Optimisation of diagnosis and treatment pathways for HCV in Tuscany – a 3-year project funded by Bando Ricerca Salute 2018 of Tuscany Region (Italy) for supporting the regional bodies in the implementation of the HCV regional plan for 2018-2020 (DGR397/2018) (ethical approval received by the Regional Ethics Committee of Tuscany Region).

We used four sources of data: (i) outpatient care data (SPA); (ii) drug prescription data (SPF and FES); (iii) exemption codes data flow (RFC 192); and (iv) data about socio-demographic information on all individuals enrolled in the Tuscan healthcare system, including sex, date of birth, and date of death.

### Data manipulation

We built a dataset that included monthly observations - collected between January 2015 and December 2022 - on i) the number of serological tests to detect HCV (SPA) ii) the number of PCR tests to detect HCV (SPA) and iii) the number of prescriptions of direct-acting antivirals (SPF and FES). Individuals who had been linked to at least one step of the hepatitis C care pathway between 2015 and 2021 were included in the study. These were individuals who had received a screening or diagnostic HCV test and/or a DAA prescription recorded in the health information system. Outpatient services data were used to identify patients that had entered the HCV pathway via screening or through diagnostic test at least once in the study period. Pharmaceutical services records gather data about drug prescriptions which were used to identify patients treated with DAA. Each patient in the dataset was identified through a unique code conferred by the regional agency for healthcare to ensure pseudo-anonymization. This aggregation was implemented while considering i) the individuals enrolled in the Tuscan healthcare system; ii) the sub-population of patients aged between 33 and 53 years old targeted by the national HCV screening; iii) sub-population at high risk for HCV infection, namely PLP and PWUD. Data on both serological and PCR tests for the diagnosis of HCV were included assuming serological test was performed for screening purposes while PCR for HCV-RNA identification was performed for diagnosis confirmation purposes and for treatment follow up. Data on the number of tests performed were sourced from SPA, however no test result was available. We used the information on the confirmatory PCR test as a proxy to identify HCV-infected individuals. We defined the primary outcome of interest as the ratio between the number of prescriptions for DAA and the number of confirmatory PCR tests conducted in the preceding month. This ratio estimated the extent of timely linkage to care and treatment for HCV-infected individuals recently diagnosed. The one-month period was defined based on available data: on average, patients who make the serological test undergo a confirmatory PCR test within one month and receive a DAA prescription within the following 4-week period.

### Interrupted Time Series Approach

To assess the effect of the regional action plan implemented by Tuscany Regional Health Authority, we used a methodological framework based on *Interrupted Time Series*. Interrupted time series analysis^25–26^ is a statistical methodology used to evaluate the impact of an intervention on a particular outcome over time. It is a very flexible framework employed to analyze empirical applications that refer to various fields of social sciences including public health^27–28^, economics^29–30^, political sciences^31–32^, and education^33–34^. The goal is to assess whether an intervention causes a significant change in the trend of a dependent variable (the outcome). In an interrupted time series design, data are collected at multiple time points both before and after the intervention. The intervention is introduced at a specific point in time, known as the *interruption*. The key idea is to compare the trajectory of the outcome variable before and after the intervention to estimate the actual effect of the intervention.

We obtained monthly observations of all the quantities of interest, observed between January 2015 and December 2022. The intervention of interest was the AIFA resolution promoted in April 2017, which was represented by a dummy variable that equals 1 in all the month-year observations occurring after April 2017, and 0 otherwise. The primary outcome of our study was given by the ratio between the number of prescriptions for direct-acting antivirals against HCV and the number of PCR tests conducted to detect HCV in the preceding month. This response variable has been normalized according to an *Ordered-Quantile (ORQ)* normalization transformation^39^.

We also included as control variables the number of serological tests to detect HCV performed two months prior to the observation and a dummy variable which equals one in all the month-year observations occurring after the identification of the first COVID cases (we considered March 2020 as starting point of the COVID pandemic). Moreover, introducing lagged structure of the data allows to account for potential endogeneity. To also account for temporal autocorrelation, we chose to adopt an *ARIMA* (AutoRegressive Integrated Moving Average)^35–36^ model specification within the ITS design. ARIMA models are particularly useful when the data shows a non-random, time-dependent structure^37^. The ARIMA model is defined by three parameters (*p, d, q)*: the parameter *p* is the order of the AutoRegressive (AR) component, which represents the number of lagged observations used for prediction; the parameter *d* is the degree of differencing (integration) required to make the time series stationary. If *d = 0*, it means no differencing is needed; if *d = 1*, first-order differencing is applied, and so on; *q* is the order of the Moving Average (MA) component, which represents the number of lagged forecast errors used for prediction. The order of these components is usually determined through data exploration and analysis: for instance, it can be chosen by examining the autocorrelation and partial autocorrelation plots or by checking at the goodness of fit and the performance of various statistical models, defined using alternative values of the parameters *(p, d, q).* In this work, we decided to follow this second approach and, for each model, we decided the value of the parameters *(p, d, q)* by comparing results obtained using different values of these parameters (the choice of the parameters results from the implementation of the *auto.arima* function of the Forecast R package in R)^38^.

## RESULTS

In this section, we mainly focus on the results obtained in the general cohort of people, in the general cohort of people aged between 33 and 53 and in the cohort that include both PLP and PWUD; however, all the other findings are available in the Supplementary Material - Appendix B.

In the observed time frame, Tuscany regional health authority delivered, 561874 serological tests, 79912 PCR tests and 13098 DAA prescriptions; people in the generic population received 561874 serological tests, 79912 PCR tests and 13098 DAA prescriptions; people aged between 33 and 53 received 242725 serological tests, 25370 PCR tests and 4695 DAA, while people with exemptions (PLP and PWUD) received 15191 serological tests, 4361 PCR tests and 1210 DAA prescriptions. In the same period, 163177 people aged between 33 and 53 received a serological test to detect early signs of HCV, 10079 individuals received a screening through PCR test and 4339 were treated through DAA. Figures A1, A2 and A3 in the Supplementary Material, Appendix A, illustrate the distribution of prescribed DAA, the number of examined PCR tests, and the number of examined serological tests over time for the general population, for the general population aged 33-53 and the population that includes PLP and PWUD, respectively. As observed, the number of tests (both PCR and serological) increased over time. Similarly, the number of prescribed DAA has increased over time.

Figures 1, 2 and 3 show the results obtained in the ARIMA model, in the general population, in the sub-population of individuals aged between 33 and 53 and in the sub-population of individuals that are either PLP or PWUD, respectively. Tables reporting all the estimates are available in the Appendix These three figures illustrate the results in the context of a counterfactual examination. In particular, the actual values of the response variable (grey dots), the predicted values of the model (blue line), the predicted values of the response variable obtained in the absence of the intervention (orange line) and the predicted values of the response variable obtained in the presence of the intervention but in the absence of the COVID pandemic (black line) are reported in these graphs. In both figures, the orange line lies below the blue line indicating that the intervention had a positive effect and improved HCV treatment coverage. Furthermore, as the black line lies above the blue line, the impact of AIFA resolution was shrunk by the spread of the COVID pandemic.

**Figure 1.**
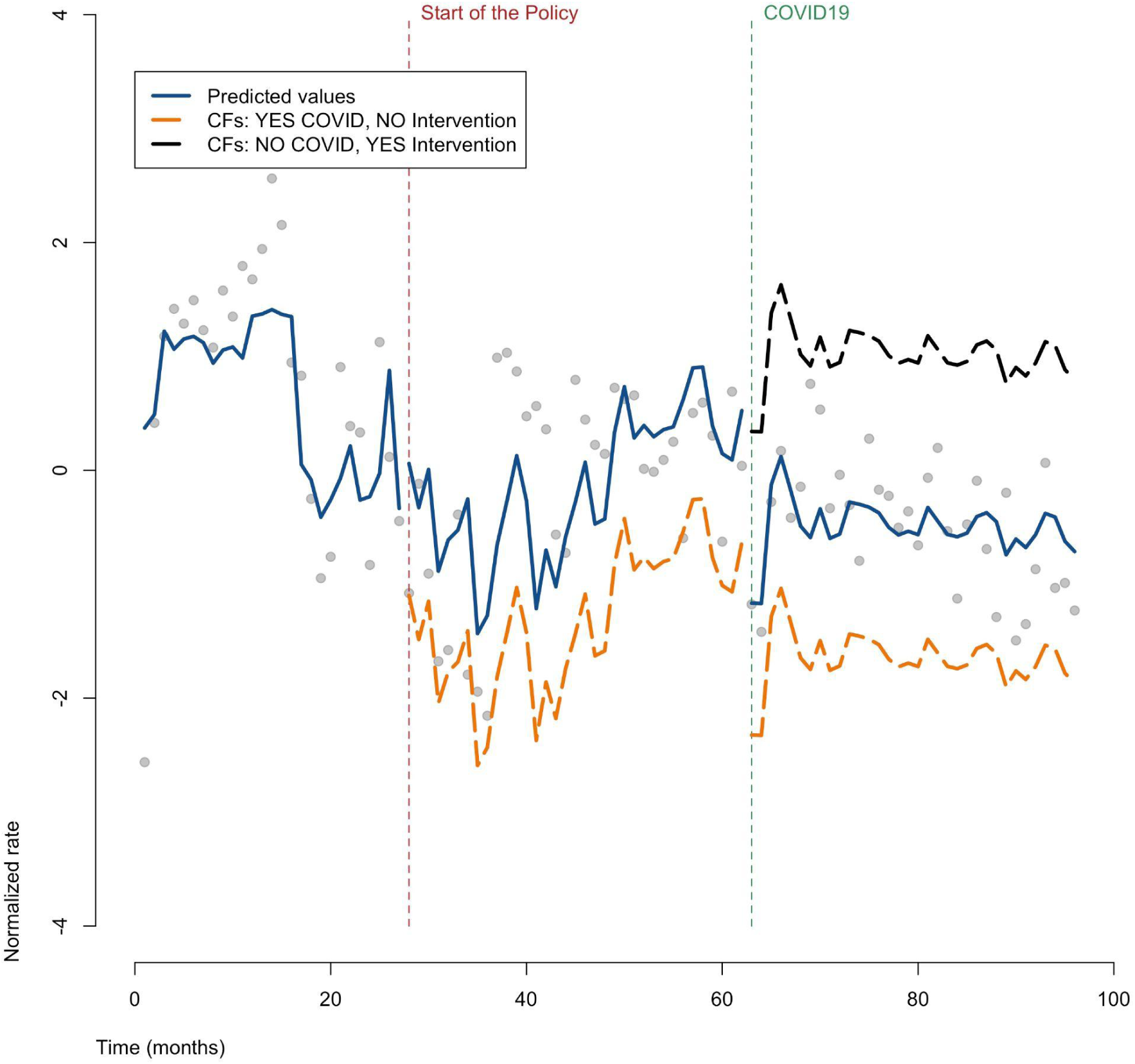
ITS, ARIMA Results. Counterfactuals. General population.

**Figure 2.**
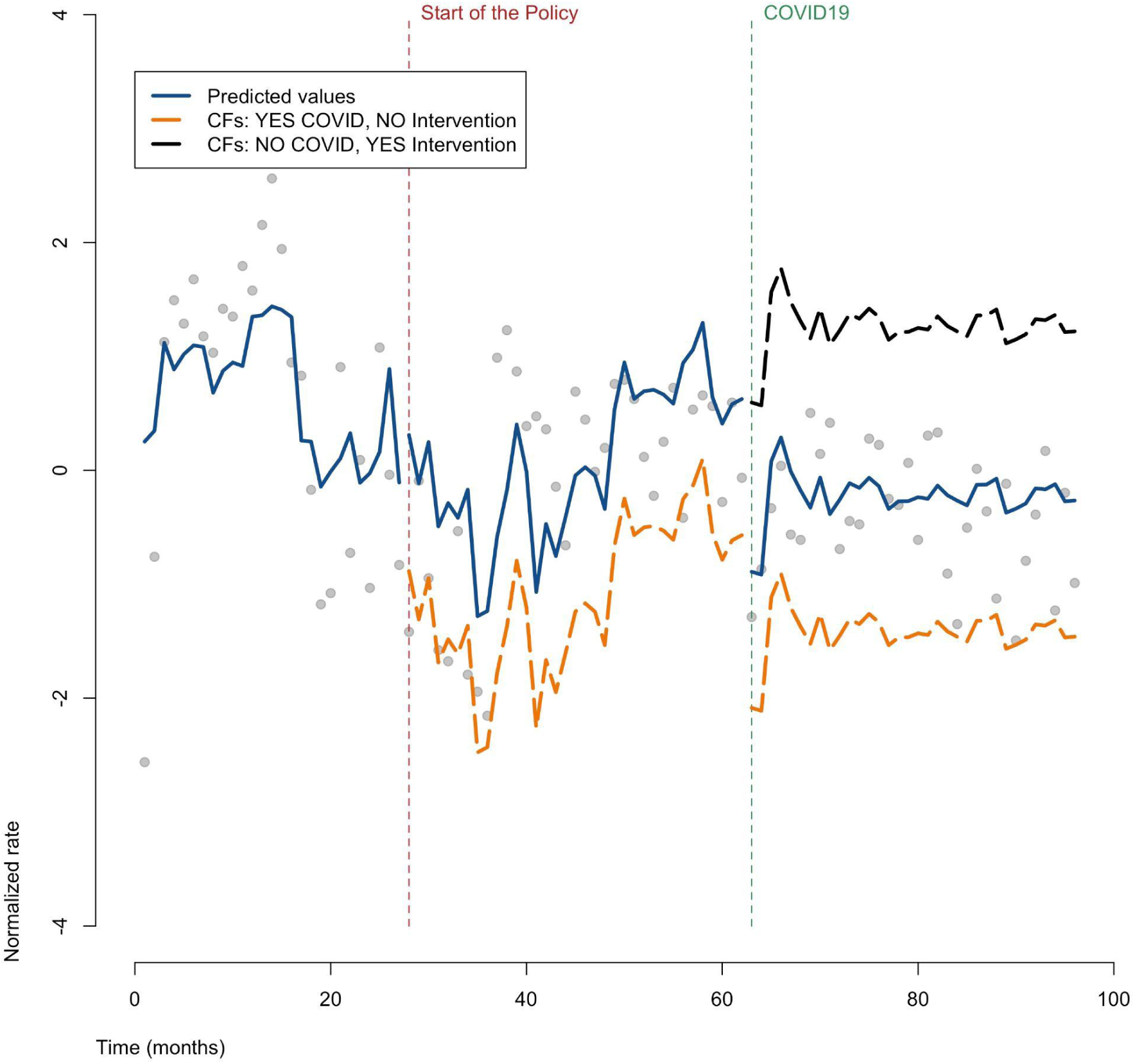
ITS, ARIMA Results. Counterfactuals. General population, people aged between 33 and 53.

**Figure 3.**
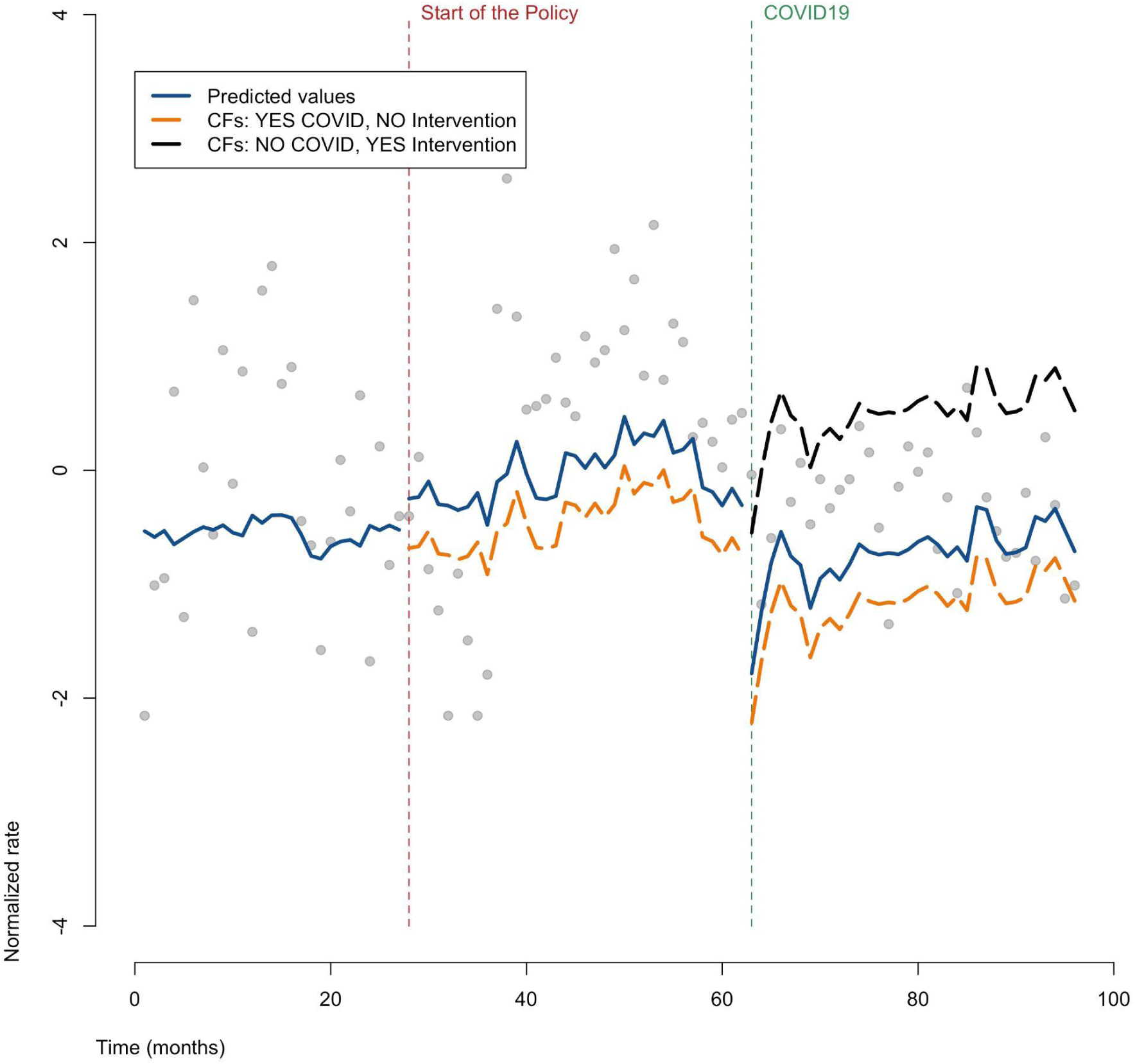
ITS, ARIMA Results. Counterfactuals. PLP and PWUD.

Finally, Figure 4 presents a comparison of the estimated ARIMA coefficients (and 95% confidence intervals) related to the effect of the intervention; Figure A4 in the Supplementary Material – Appendix A shows the effect of the COVID pandemic for all identified sub-populations.

**Figure 4.**
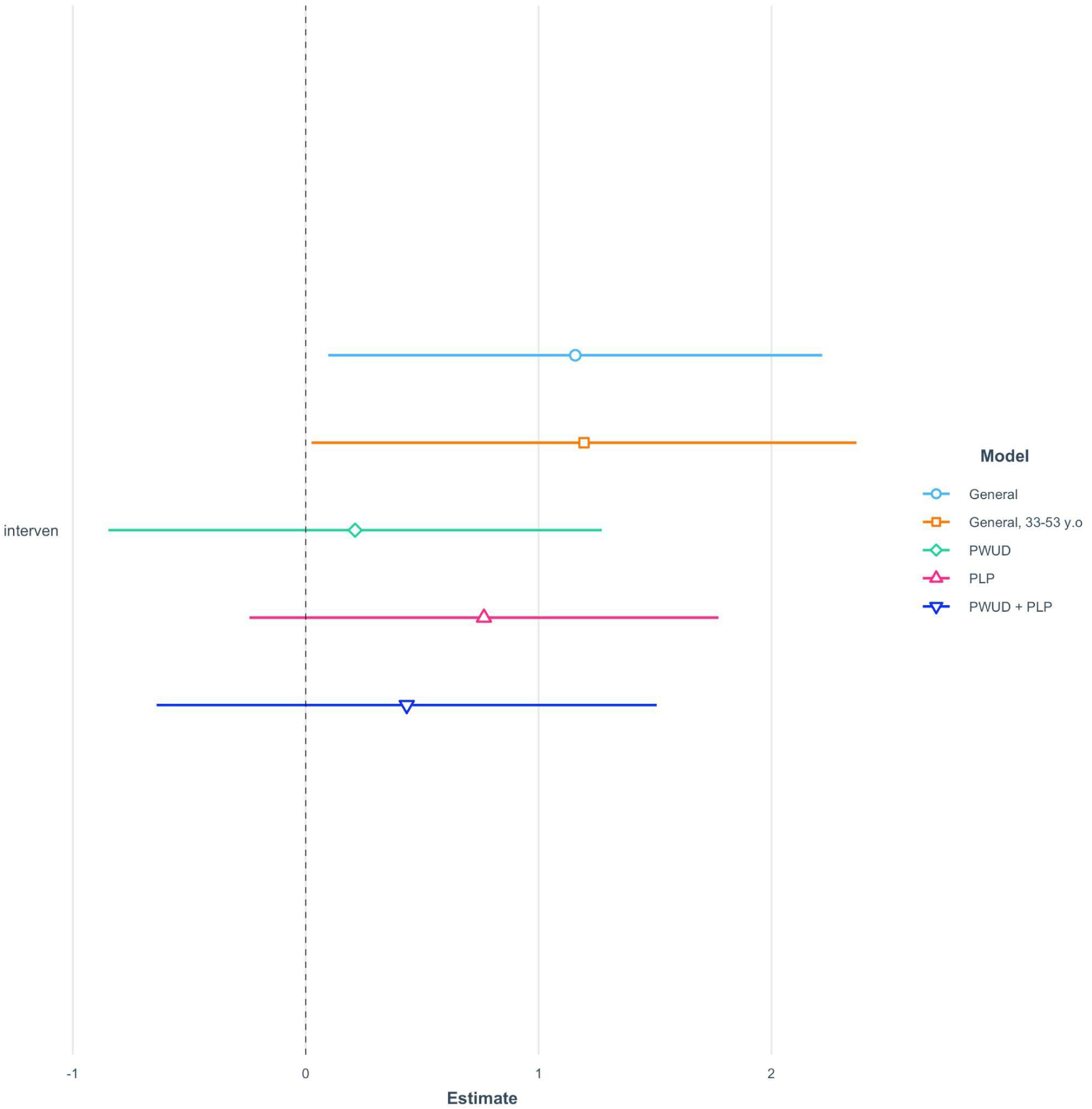
ITS, ARIMA Results. Comparison of coefficients: the effect of the intervention.

As can be observed, the effect of the intervention was positive and statistically significant for the general population and selected ages. However, the effect was non-significant for PLP and PWUD (figure 4). On the contrary, COVID-19 had a negative impact on all population groups, except for PLP for whom non-significant effect was observed (figure A4 Supplementary Material – Appendix A).

## DISCUSSION

Here we attempt to give an interpretation to the data obtained through an interrupted time series analysis used to assess the impact of the policies of the Region of Tuscany, Italy, promulgated to achieve the goals of HCV elimination as identified by the WHO.

Firstly, it is important to highlight the effect of relaxing the criteria for access to DAA treatment on HCV diagnostic and therapeutic performance. Indeed, the 2017 AIFA resolution on extended access to DAA made access to treatment universalistic, allowing all people living with HCV to start DAA therapy regardless of the stage of liver fibrosis. This indicates that the Tuscany regional health authority raised their awareness of the importance of preventing HCV, resulting in a progressive effort to enhance screening and, consequently, the increase in initiated treatments with DAA. However, with the passage of time, after the first wave of new treatment initiations, expanding access did not result in sustained increases in the search for new cases through continuous and targeted screening, as Herink and colleagues also pointed out in their study^40^. In fact, retesting of patients already linked to care, prior to DAA treatment initiation may have contributed to the observed initial peak in screening.

A remark must be made about the high number of tests provided to women during the study period. Even before 2015, in Tuscany Region as well as in the national territory, a provision was in force - and still is - to test for HCV antibodies during the first trimester of pregnancy. This results in a high flow of tests provided to women of childbearing age, partly overlapping with the 1969-1989 birth cohort considered in the study. However, this screening effort does not correspond to a high number of HCV treatments prescribed to women, in line with other studies reporting a low prevalence of HCV among pregnant women^41^.

Parallel to the increase in access to diagnostic services, which led to an increased number of new diagnoses, the number of patients initiated on treatment also saw a marked increase, particularly between 2017 and 2018. This spike in the provision of DAA treatment in the first year after the AIFA resolution can be explained by the fact that all patients already diagnosed and linked to care were recalled to start treatment. The resolution had a positive and substantial effect on the increase in the number of treatments provided because of the broadening of the inclusion criteria leading to universal access. In addition the Regional Health Authority implemented the ministerial indications and endorsed a regional HCV elimination plans, thereby favoring the linkage to care of people living with HCV infection. As the work of Kondili and colleagues shows, the effect of the national resolution on regional policies led Italy to be one of the twelve nations on course to meet the WHO’s 2030 HCV elimination targets, provided that 40 000 patients receive treatment year until 2030^42^. This trend was also confirmed in other studies^43^.

As our results show, the effect of the intervention was positive and statistically significant in the general population and the selected birth cohort. However, this effect becomes statistically non-significant in sub-populations consisting of PLP or PWUD. This finding could be explained with the fact that both PLP and PWUD are well characterized for their heightened prevalence of HCV, and they may have been targeted for HCV screening and treatment efforts before 2017. Although PWUD and PLP may be engaged in screening programs via dedicated healthcare services, namely harm reduction services or prison healthcare services, scarce evidence is available at local and national level as to the extent of such interventions in recent years^14^. On the other hand, these results could be explained by a scarce effort of public health authorities in targeting these specific populations before and after the interventions, despite robust knowledge about drug use as a risk factor, as also demonstrated in modeling studies^44^.

In order to tackle this and to promote the identification of undiagnosed cases, especially among population groups where virus transmission is most frequent, a region-wide screening campaign was launched in 2023. Already since the end of 2022 in harm reduction services (SerDs) and prisons, an intensified screening effort was launched. However, it is likely that the rate of acceptance and initiation of treatment is lower in these settings, partly due to higher rate of treatment discontinuation (e.g. post-release discontinuity of care) or suboptimal compliance, and partly due to the challenges in reaching PWUDs who are not linked to harm reduction services.

In the general population, the triennial HCV elimination action plan promoted by the Tuscany Regional Health Authority promoted access to DAA and increased the coverage of HCV treatment, as demonstrated previously. This effect would have been even more pronounced if the COVID pandemic had not affected the progress starting from March 2020. The COVID pandemic had a negative and significant effect on the general population and on all examined sub-populations, resulting in a reduced number of HCV treatment initiations . This evidence has been brought to light by a number of other studies^45–46^. In particular, the work of Binka and colleagues^47^, through an interrupted time series analysis, shows how public health measures related to COVID-19 had a negative effect on HCV screening and diagnosis performance, reducing the number of HCV diagnoses in Canada. In parallel and as a consequence of this, the number of treatments delivered also declined: global trends in DAA utilisation have been explored extensively in a number of studies, which concluded on a negative impact of the COVID-19 pandemic^48^. Konstantelos and colleagues have also shed light on the same phenomenon in Ontario, Canada, obtaining results overlapping with those emerging from our work in a high-income country with universalist health policies similar to Italy^49^. Because of the COVID-19 pandemic, as well as of other intervening factors, Tuscany Regional health authority failed to meet the targets for treatment set in the 2018 regional action plan, in terms both of new treatments initiated and new diagnosis.

Our research has some limitations. First of all, information on tests carried out by private providers (such as non-governmental organizations) is not included in administrative health records used to determine outpatient services and pharmaceutical services. Additionally, information on the outcomes of laboratory tests and treatments could not be retrieved. We were unable to evaluate the development of the HCV continuum of care because the laboratory test results of the patients were unavailable. Yousafzai et al.^50^ documented the HCV care cascade following the introduction of DAAs in Australia in a recent study utilizing the same methodological technique by accessing HCV test results through data linkage. Second, we gathered data on treatment prescriptions; however, since data on patients’ actual drug use was not available, we used prescriptions as a stand-in for DAA uptake, even if we do not have information about the treatments’ outcomes.

In order to, link data, we identified PWUD using exemption numbers. Though not particularly for intravenous drugs, the same code is linked to all substance use disorders, which likely causes an overestimation of PWUD in our study. However, acquiring an exemption code from the SSR requires linkage to care (such as HR services), which could result in an underestimate of the real PWUD population living in Tuscany throughout the study period.

Additionally, testing services given prior to 2015 were not considered because our analysis began with the introduction of DAAs in that year. This might have caused the proportion of the Tuscan population who had undergone HCV screening at least once to be underestimated.

## CONCLUSIONS

The results of this study indicate a significant upsurge in activities associated with the screening of Hepatitis C Virus (HCV) in Tuscany between 2015 and 2022. Furthermore, it is evident that the triennial action plan implemented by the regional authorities following the 2017 AIFA resolution has substantially enlarged the treatment coverage of HCV in the general population and that the effect of the policy would have been even stronger if the COVID pandemic had not affected service provision since March 2020. In the sub-population of PLPs and PWUDs the effect of the intervention was not statistically significant, suggesting that treatment coverage remained substantially stable after the implementation of the policies. To conclude, the strategic policy implemented by the Tuscany Regional Health Authority following the AIFA resolution to enhance DAAs coverage has proven to be remarkably effective in the general population, while additional efforts may be needed to target equally effectively population sub-groups such as PLPs and PWUDs. Through a targeted approach, the region has succeeded in expanding access to DAA prescriptions, demonstrating a clear commitment to combating Hepatitis C.

The findings presented in this contribution serve as compelling evidence, urging other national and subnational authorities to enhance HCV and streamline access to DAA prescriptions. In the years immediately following the AIFA resolution, the regional plan of the Tuscany Region Health Authority had an important effect in increasing the distribution of the treatment. However, it has not been as effective in actively proposing screening for population groups at high risk of transmission, nor for the general population. This is the context of the national screening program, to which Tuscany Region has adhered, that will start in full force and effect in mid-2023. The role of the screening campaign is crucial for several compelling reasons. First, timely screening enables the early detection of HCV infections, facilitating swift intervention and treatment initiation. Identifying cases in the early stages is critical to prevent disease progression and reduce the risk of severe complications. Second, promptly detecting individuals with HCV curbs the potential for further transmission within communities. Lastly, from a public health perspective, widespread screening aids in comprehending the virus’s prevalence in different communities, guiding targeted interventions and resource allocation for effective disease management.

In addition to the screening campaign, it is also crucial to facilitate access to DAA prescriptions, to enhance treatment accessibility. Simplifying the procedures for healthcare providers to prescribe these antivirals for free ensures that affected individuals, regardless of their socio-economic status or geographic location, can readily access and benefit from modern, effective treatments. From a public health perspective, early detection and treatment of HCV can lead to substantial healthcare cost savings in the long run. Indeed, by preventing the progression of the disease to advanced stages, authorities can reduce the economic burden associated with treating severe complications and improve the efficiency of the whole healthcare system.

From a broader perspective, increased screening and improved access to DAA prescriptions align with the worldwide efforts to eliminate HCV as a public health threat. National and subnational authorities play a pivotal role in contributing to international goals by implementing effective screening programs and ensuring that appropriate treatments are readily available. In conclusion, findings of this study provide evidence to support policies at national and subnational levels to booster HCV screening and simplify access to DAA prescriptions. This not only enhances individual health outcomes but also holds broader implications for public health, disease prevention, and global initiatives to eliminate HCV.

## Data Availability

All relevant data are within the manuscript and its Supporting Information files

## Acknowledgments

This study was supported by the regional administration - Direzione Sanità, Welfare e Coesione sociale – of Tuscany (Italy). The authors would like to thank the management and healthcare laboratory of the Sant’Anna School of Advances Studies researchers in the person of Professor Milena Vainieri for the chance to use data from Administrative Healthcare Records. More detailed data regarding the effectiveness of the screening campaign will be available in the future to update this work.

## Notes

### Competing Interest Statement

The authors have declared no competing interest.

### Funding Statement

The author(s) received no specific funding for this work.

### Author Declarations

Ethical approval received by the Regional Ethics Committee of Tuscany Region

